# Biometric covariates and outcome in COVID-19 patients: Are we looking close enough?

**DOI:** 10.1101/2020.11.04.20225961

**Authors:** Sebastian Fritsch, Konstantin Sharafutdinov, Gernot Marx, Andreas Schuppert, Johannes Bickenbach

## Abstract

**Background:** The impact of biometric covariates on risk for adverse outcomes of COVID-19 disease was assessed by numerous observational studies on unstratified cohorts, which show great heterogeneity. However, multilevel evaluations to find possible complex, e. g. non-monotonic multi-variate patterns reflecting mutual interference of parameters are missing. We used a more detailed, computational analysis to investigate the influence of biometric differences on mortality and disease evolution among severely ill COVID-19 patients.

**Methods:** We analyzed a group of COVID-19 patients requiring Intensive care unit (ICU) treatment. For further analysis, the study group was segmented into six subgroups according to BMI and age. To link the BMI/age derived subgroups with risk factors, we performed an enrichment analysis of diagnostic parameters and comorbidities. To suppress spurious patterns, multiple segmentations were analyzed and integrated into a consensus score for each analysis step.

**Results:** We analyzed 81 COVID-19 patients, of whom 67 required MV. Mean mortality was 35.8 %. We found a complex, non-monotonic interaction between age, BMI and mortality. A subcohort of patients with younger age and intermediate BMI exhibited a strongly reduced mortality risk (p < 0.001), while differences in all other groups were not significant. Univariate impacts of BMI or age on mortality were missing. Comparing MV with non-MV patients, we found an enrichment of baseline CRP, PCT and D-Dimers within the MV-group, but not when comparing survivors vs. non-survivors within the MV patient group.

**Conclusions:** The aim of this study was to get a more detailed insight into the influence of biometric covariates on the outcome of COVID-19 patients with high degree of severity. We found that survival in MV is affected by complex interactions of covariates differing to the reported covariates, which are hidden in generic, non-stratified studies on risk factors. Hence, our study suggests that a detailed, multivariate pattern analysis on larger patient cohorts reflecting the specific disease stages might reveal more specific patterns of risk factors supporting individually adapted treatment strategies.

## Background

The novel Severe acute respiratory syndrome Coronavirus 2 (SARS-CoV2)-infection COVID-19 most commonly presents with mild symptoms like fever, malaise and symptoms of an upper respiratory tract infection (1). Among German patients, hospitalization is necessary only in about 14 % of cases (2). Those in need of inhouse treatment with COVID-19 can often be handled at general wards while only a minority of patients with a fulminant deterioration is in need for intensive care resources and consecutive ventilatory support (3). However, this particular patient group that most predominantly presents with Acute respiratory distress syndrome (ARDS) and severe hypoxia requires complex and extensive treatment including timely endotracheal intubation and lung protective concepts of invasive mechanical ventilation (MV), high PEEP levels, inhalative nitrous oxide, proning, and, in case of refractory hypoxia, ECMO therapy (4).

For a targeted use of the available intensive care beds, it is of great importance to know which patients are particularly at risk of suffering a severe course. The impact of biometric covariates on risk for severe outcomes of COVID-19 disease was assessed by numerous observational studies on unstratified cohorts (5-8). Several international publications identified a higher age, male gender and an increasing number of comorbidities as risk factors for a poor outcome (9, 10). However, within the existing studies, there is great heterogeneity concerning the estimated influence of single biometrical parameters as well as differing clinical outcomes themselves. Terms of hospitalization and Intensive care unit (ICU) resources differ as well (5, 7, 8, 11, 12), which makes a reasonable comparison between the studies difficult. Possible reasons for different outcomes may lie in large heterogeneities of health care systems, hospital and especially ICU resources as well as differing admission policies and clinical operating instructions. Another obstacle for a clear picture is the considerably differing population under analysis between the studies. Some studies included only hospitalized and deceased patients, while in other publications, non-hospitalized patients with milder courses of disease were included, which led to different estimations for the impact of biometric covariates, as well. However, multilevel evaluations, which are able to find possible complex, e. g. non-monotonic multi-variate patterns reflecting mutual interference of parameters, are usually missing. The fact that there is a lack of clarity about the real significance of certain factors has led to uncertainty and partial rejection of prophylactic protective measures in parts of the public (13). To elucidate the impact of different biometric risk factors on the course of the novel disease, it might be necessary to go into a more detailed, computational analysis.

To give an example of these more advanced techniques, we performed a retrospective analysis of a dataset of COVID-19 patients. Within this study, we therefore aimed to investigate the influence of biometric differences on mortality and disease evolution within a cohort of severely ill COVID-19 patients.

## Methods

This retrospective analysis included patients with a confirmed diagnosis of COVID-19 between March and May 2020, who were admitted to our University Hospital RWTH Aachen (UKA) and needed treatment on an Intensive Care Unit (ICU). All patients had a positive result on a SARS-CoV2-PCR assay of a specimen collected on a nasopharyngeal swab.

This analysis was approved by the local ethical review board (EK 091/20; Ethics Committee, Faculty of Medicine, RWTH Aachen, Aachen, Germany). Patients’ confidentiality was maintained.

Data were retrieved from an electronic patient data recording system (medico//s, Siemens, Germany) and from an online patient data documentary system (IntelliSpace Critical Care and Anesthesia, ICCA Rev. F.01.01.001, Philips Electronics, The Netherlands). All data from patients admitted to the ICUs were included and merged into a data register (Excel, Version 16.37, Microsoft Corporation, Redmond, WA, USA).

Acute respiratory distress syndrome (ARDS) and its severity was classified according to the grades of hypoxia as defined by the so called “Berlin definition”.

The recorded data contained biometric parameters like age and gender, preexisting comorbidities and a set of vital signs, laboratory parameters and the ventilator settings. For a full list, refer to the Supplement. Mortality was chosen as primary endpoint, while length of MV represented a secondary endpoint.

Since age and body mass index (BMI) were described previously as relevant for the evolution of disease, they were particularly chosen for a more detailed analysis. For the analysis of the interactions between BMI, age and mortality, the population was divided into three BMI subgroups, defined as low, intermediate and high BMI group and a lower and a higher age group. In order to minimize spurious effects from subgroup splitting, we analyzed subgroups across a bundle of subgroup settings. The respective splitting value were 25 and 27 for the lower and 30 and 32 for the upper limit. Age splitting was carried out at the bounds 65, 68 and 70 years. These splitting resulted in 12 possible combinations of subgroups, which were analyzed individually. To avoid very small subgroups, no further splitting of the BMI-age defined subgroups with respect to gender was carried out. The 12 possible combinations, which resulted from applying variable subgroup borders, were summarized in 6 subcohorts by calculating a mean value for the different groups with a combination from BMI (low/intermediate/high) and age (low/high). By that, outcomes and identification of diagnostic parameters, which are specific for each of the 6 subgroups, were calculated.

To identify risk factors associated with the diverse outcome across the biometric subcohorts, we calculated differential expression for preexisting comorbidities and for 54 diagnostic, baseline value parameters assessed in the ICU, i.e. the first available measurements of respective parameters.

All statistical analyses were performed using Matlab R2015b, Statistical and Machine Learning Toolbox (The MathWorks, Inc.).

Primarily, the impact of BMI and age on mortality was analyzed. Second, the distribution of age and BMI in survivors and non-survivors was further calculated individually (Wilcoxon-Ranksum test). Based on these results, we stratified BMI into three subgroups as described above and analyzed the age distribution of survivors and non-survivors in each subcohort. The same analysis was performed for length of MV as secondary endpoint, followed by statistical analysis of relation between the two endpoints. Finally, analogous calculation was performed comparing a cohort of non-ventilated ICU patients with a ventilated cohort. To complete the analysis, we analyzed the enrichment of comorbidities as well as differential expression of the baseline diagnostic parameters between MV survivors with length of mechanical ventilation below 30 days and more than 30 days, respectively.

For each subcohort we performed enrichment of mortality as well using hypergeometric cumulative distribution testing (Matlab: hygecdf).

## Results

### Patient characteristics

We analyzed data of 81 COVID-19 patients, of whom 67 required MV during their ICU stay. Clinical characteristics of the complete population are shown in Table 1.

**Table 1.** Clinical characteristics of the analyzes ICU patient cohort

|  |  |
| --- | --- |
| Total number of patients, n | 81 |
| Age (mean ±SD), years | 64.3 ± 11.3 |
| Male gender, n (%) | 54 (66.6 %) |
| Length of stay ICU, days (mean ±SD) | 25.8 ± 22.0 |
| MV, n (%) | 67 (83 %) |
| Length of MV, days (mean ±SD) | 26.5 ± 20.6 |
| ARDS, n (%) | 57 (70.4 %) |
| - Mild, n (%) | 1 (1.2 %) |
| - Moderate, n (%) | 21 (25.9 %) |
| - Severe (n; %) | 35 (43.2 %) |
| Pulmonary Embolism (n; %) | 17 (21.0 %) |
| Renal replacement therapy (n; %) | 46 (56.8 %) |
| ECMO (n; %) | 16 (19.8 %) |
| Mortality (n; %) | 29 (35.8 %) |

### Influence of BMI and age on mortality and length of MV

Data distribution across BMI and age revealed an apparent inhomogeneity of mortality and length of mechanical ventilation across the BMI-age plane indicating a complex, non-monotonic interaction between age, BMI and mortality, as can be seen in figures 1a and 1b.

**Figure 1a:**
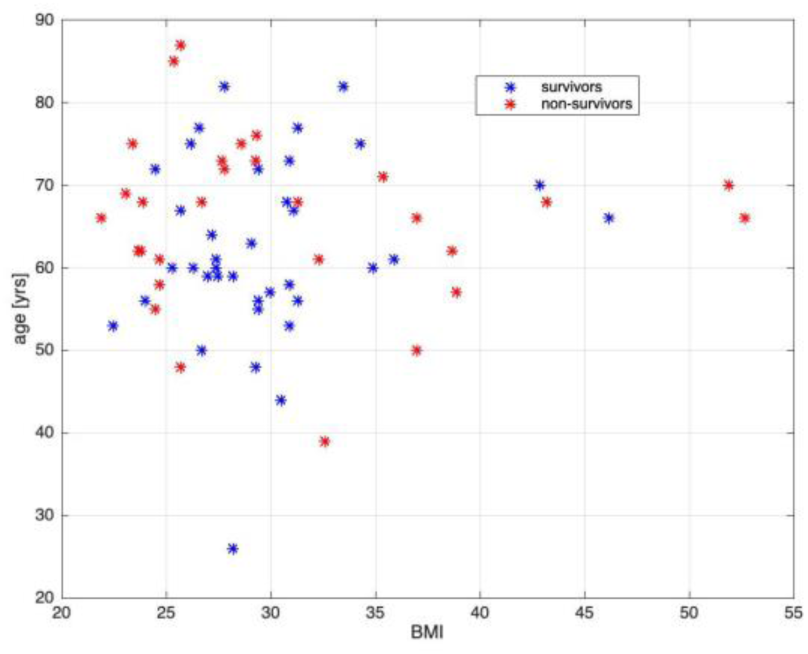
Distribution of mortality across combined BMI – age data in COVID-19 ICU patient cohort

**Figure 1b:**
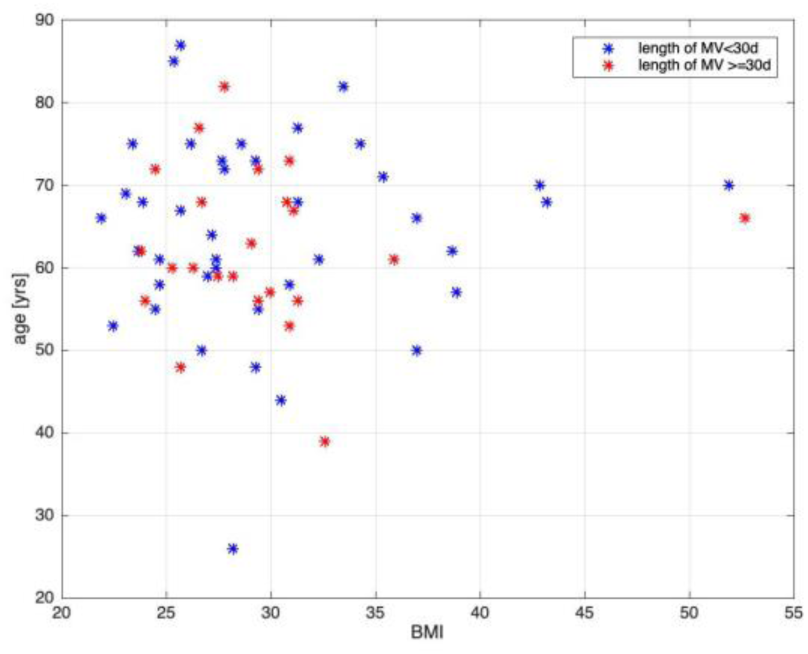
Distribution of length of MV across combined BMI – age data in COVID-19 ICU patient cohort

For both BMI and age, there were no significant differences between survivors and non-survivors (p = 0.65 for BMI, p = 0.098 for age), indicating no significant univariate impact of both BMI and age on mortality in our cohort.

As described above, patients were stratified into three BMI subgroups (low, intermediate, high). Analyzing the age distributions of survivors and non-survivors for each of these three sub-cohorts (figure 2) we found an apparent non-uniform impact of age on mortality. For the low BMI subcohort, mean age differs significantly (p = 0.025), whereas the differences of mean age for the medium and high BMI subcohorts were not significant (p = 0.29, p = 0.69).

**Figure 2:**
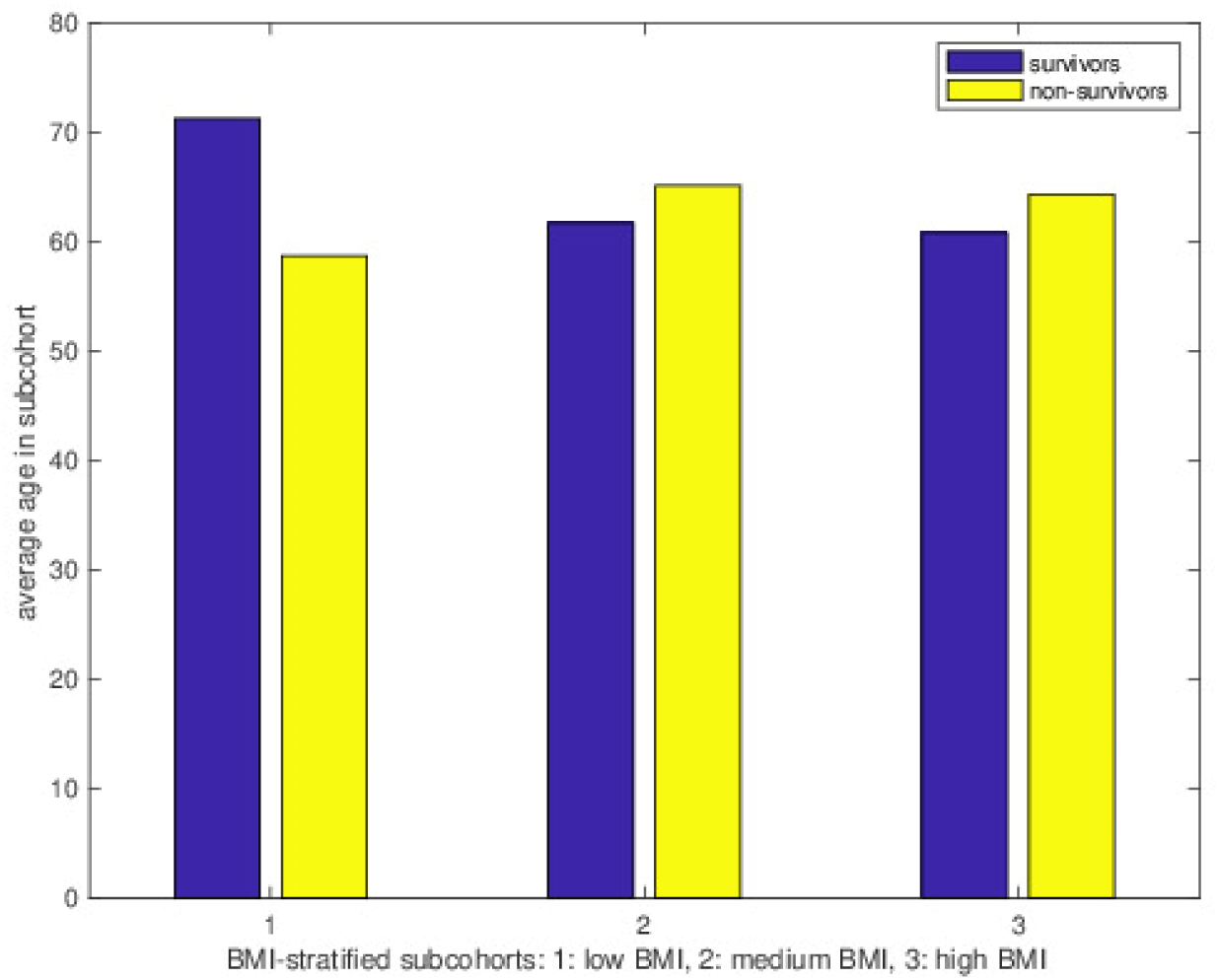
Distribution of age for survivors/non-survivors stratified by BMI

To analyze the influence of different of BMI and age in varying combinations, we generated combined BMI–age stratified subgroups as described above, resulting in 6 subcohorts. Due to the variable limits of the two parameters, the sizes of the respective populations vary within each subcohort. The average numbers are given in table 2.

**Table 2.**
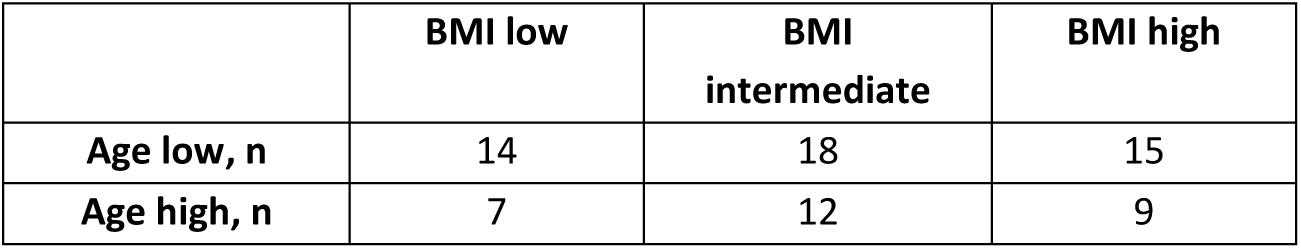
Mean number of patients in the six subcohorts. As sizes of subcohorts depend on the splitting, they are not independent explaining that the sum over the (univariate) means exceeds the number of patients

|  | BMI low | BMI intermediate | BMI high |
| --- | --- | --- | --- |
| Age low, n | 14 | 18 | 15 |
| Age high, n | 7 | 12 | 9 |

Also, after stratification in combined BMI/age groups, as depicted in figures 3a and 3b, the mortality rates reveal a complex pattern. Whereas a group of younger patients with an intermediate BMI have a very significantly reduced risk of mortality (mean risk p < 0.001), older patients, whose BMI lays within the same range, show a mean mortality. However, the impact of age is apparently inversed in high BMI groups: younger patients seem to have a slightly increased risk, whereas elderly patients have a tendency towards a decreased risk (mean risk p = 0.14) compared to the overall population. Moreover, mortality in the younger patient groups is significantly (p < 0.05) increased in 4 out of 12 splitting combinations.

**Figure 3a/b:**
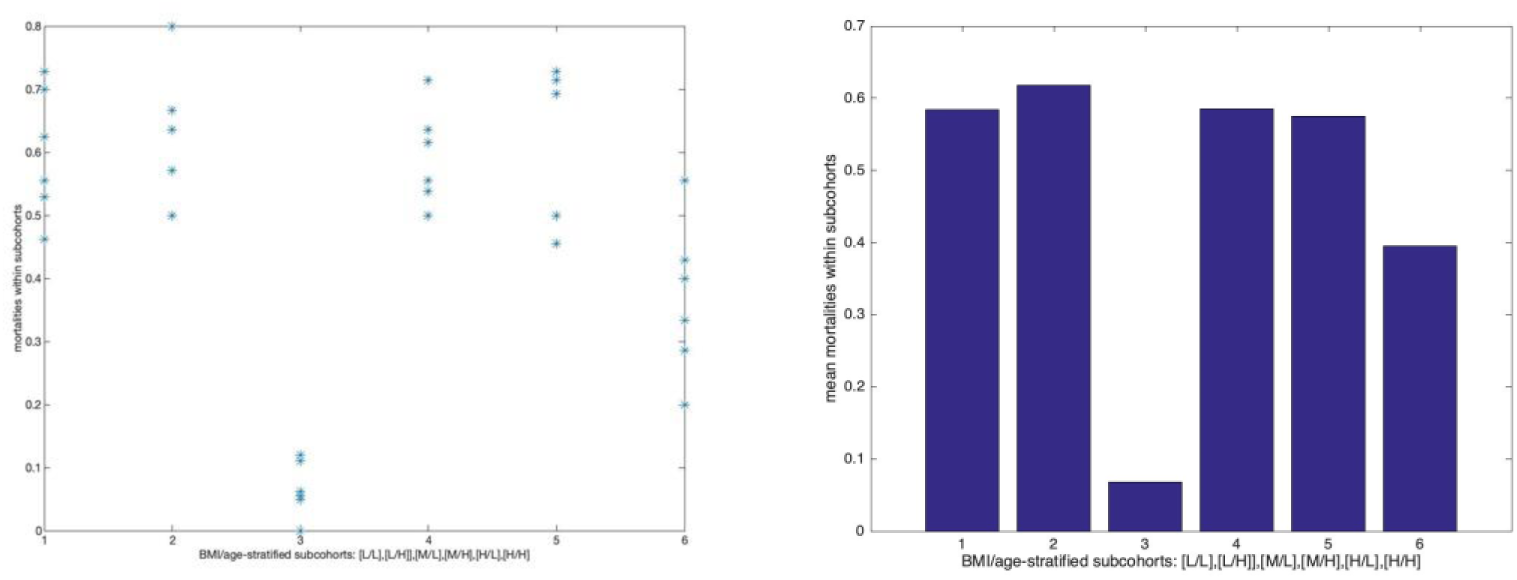
Mortality in BMI/age stratified subcohorts at 12 splitting levels reveals inhomogeneous, but non-monotonic distribution of mortality (3a). Mean mortality (weighted by subcohort size) indicate outliers of very low mortality in subcohort 3 and a slightly decreased mortality in subcohort 6. Labeling for BMI and age group: [L]: low, [M]: intermediate, [H]: high.

In contrast to our findings for mortality, the respective assessment for length of MV as secondary endpoint did not reveal significant deviations within any of the subgroups discussed above (p > 0.12). However, there is a coherent pattern for length of MV between the groups of survivors and non-survivors, as shown in figure 4. Except for the intermediate BMI/low age group, which stands out through its very low mortality, in all other groups the mean length of MV is overall significantly longer in survivors than in non-survivors. Here as well, the significance level between survivors and non-survivors in the intermediate BMI/low age group is not reached due to the low average number of non-survivors (N_average_ = 1.16).

**Figure 4:**
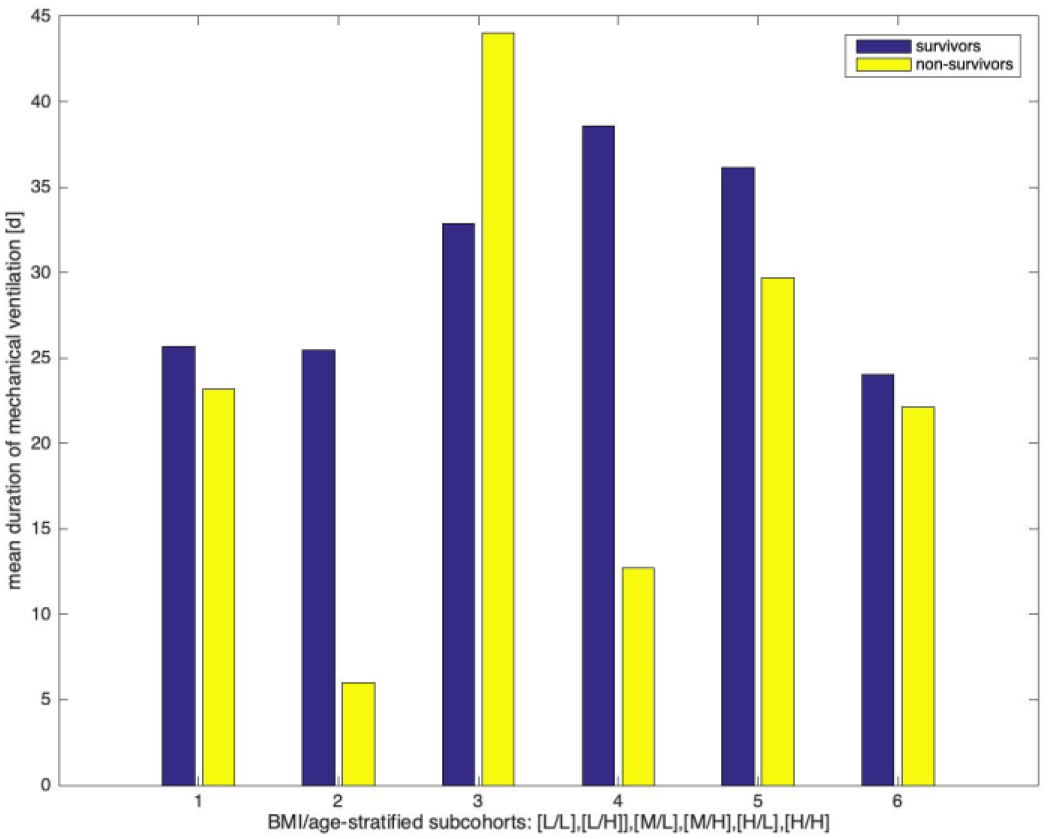
Length of mechanical ventilation is higher for survivors than for non-survivors in all subcohorts except cohort 3 (very low incidence of non-survival)

### The influence of baseline values and comorbidities

To find possible explanations for the inhomogeneous mortality rates, we aimed to examine potential differences within the baseline values, i. e. the first available measurements of diagnostic parameters. There were some significant deviations for some baseline values in single subgroups, but they did not contribute to an explanation in a reasonable way.

In analogy to baseline values, we assessed enrichment of comorbidities between survivors / non-survivors for each subcohort compared to survivors / non-survivors in all MV patients. For the entire findings, please refer to the Supplement. As the enrichment analysis did not result in neither highly significant enrichments of comorbidities nor difference in diagnostic baseline levels, corrections with respect to multiple testing showed no robust significance. Hence, we cannot claim univariate explanations of the observed patterns in mortality.

## Differences between MV and non-MV patients

COVID-19 patients that require MV represent a population with the most critical course of disease. Nevertheless, not all COVID-19 patients, who are admitted to the ICU require MV. So, to further evaluate the observed deviations of morbidity-related patterns in other epidemiological studies and our findings in mechanically ventilated ICU patients, we analyzed the diagnostic baseline parameters between overall non-MV ICU patients on the one side and all MV ICU patients / survivors and all MV ICU patients / non-survivors on the other side for significant differences.

We found two baseline-parameters exhibiting significant (Bonferroni-corrected p-values < 0.05) deviations between non-MV and MV patients. Baseline CRP is enriched in MV survivors (p < 0.001) and in MV non-survivors (p < 0.01) compared to non-MV patients. Similarly, baseline PCT is enriched in in MV survivors (p < 0.05) and in MV non-survivors (p < 0.0001) compared to non-MV patients.

Baseline D-Dimers were enriched in MV non-survivors (p < 0.01) and survivors (p < 0.001) compared to non-MV patients as well, but the p-values slightly failed the Bonferroni corrected significance level of 0.05, although giving a clear trend.

Moreover, we found a high correlation between the log10(p)-values of non-MV vs. survivors MV and non-survivors MV across all parameters (figure 5a), but no correlation between non-MV / survivor-MV and survivors-MV / non-survivors-MV (figure 5b).

**Figure 5a/b:**
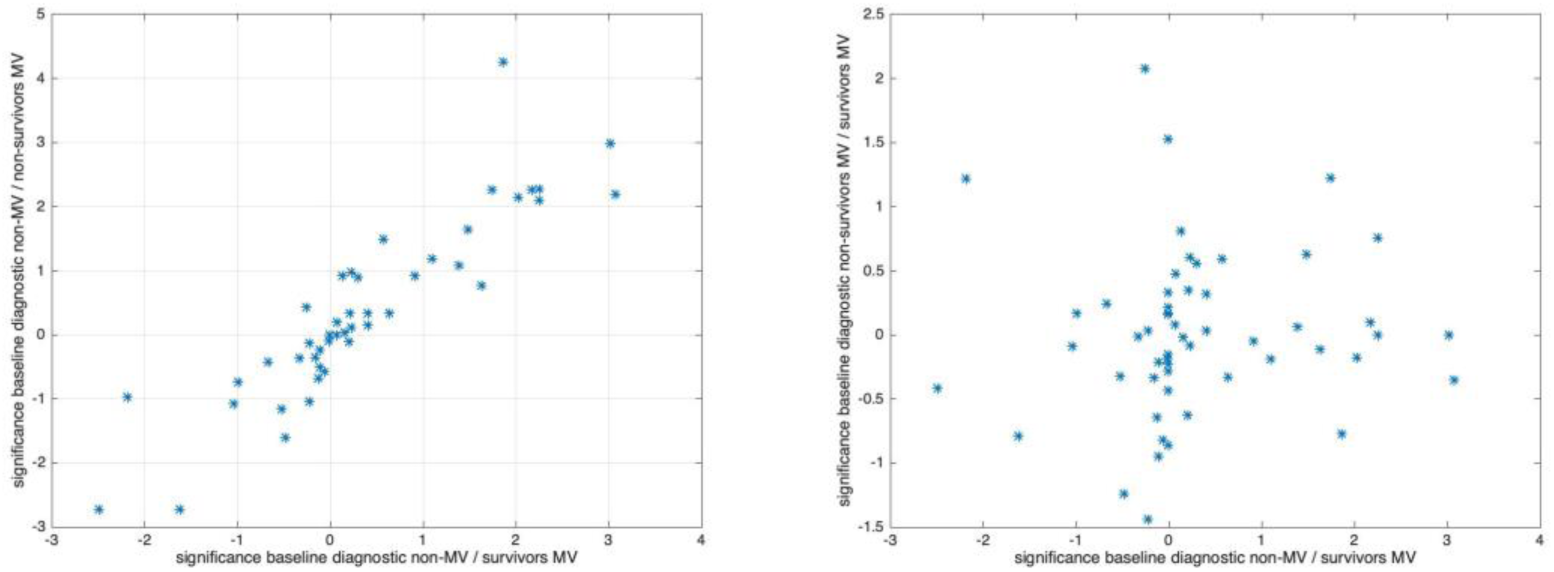
There is a high correlation between significance levels of baseline diagnostic parameters between non-MV and MV patients (5a), but no correlation when comparing survivors and non-survivors in MV (5b).

Assessing the enrichment (Bonferroni-corrected p < 0.01) of comorbidities in MV-survivor / non-survivor population compared to non-MV population, we found comorbidities, which are strongly enriched in MV cohort (independent from survival) compared to non-MV patient cohort (see Supplement). Apparently, Sepsis is highly significant enriched in MV-patient cohort compared to non-MV cohorts, both for survivors and non-survivors. Analyzing the distribution of enrichment of comorbidities in MV versus non-MV patients, we found patterns with a high similarity to the pattern received from baseline values. Similar baseline diagnostic parameters depicted in Figure 5, the log10(p)-values of enrichment of comorbidities in MV-survivors compared to non-MV and MV-non-survivors to non-MV are highly correlated (r = 0.89), whereas the respective log10(p)-values for MV-survivors vs. MV-non survivors showed no correlation (r = 0.42).

With respect to the length of MV, we found that D-Dimers were significantly higher expressed (Bonferroni corrected p-value < 0.05) in MV-survivors with more than 30 days of MV compared to less than 30 days and ‘Diseases of the blood and blood forming organs’ significantly enriched in the high length of stay subcohort as well. Both parameters show significant over-expression / enrichment in MV-survivors compared to non-MV patients as well, although the overall patterns of log10(p)-distributions show only modest correlation between both endpoints (r = 0.44, 0.42). In contrast, other baseline parameters or comorbidities controlling survival / non-survival within the MV subcohort show no correlation to duration of MV (r = 0.19, 0.36).

## Discussion

The aim of this study was to get a more detailed insight into the influence of biometric covariates on the outcome of COVID-19 patients. A relevant problem within this field is the heterogeneity of study designs and strongly differing populations leading to inconsistent results. The common coincidence of several risk factors disguises a clear estimation of their impacts on disease evolution as well. A deeper computational analysis of a larger dataset of COVID-19 patients can overcome these problems analyzing the influence of a single parameter and separating it from other accompanying parameters. To show the potentials of these systems we analyzed a dataset for the influence of biometric covariates.

In our study, we focused on patients admitted to University Hospital RWTH Aachen (UKA), who required ICU treatment. While other studies are frequently merged with mild disease stages, our study covered only patients with a highly severe health condition. In contrast to the results reported before, we found a non-monotonic impact of BMI and age on mortality and length of MV, while a univariate effect could not be determined. Furthermore, we found, that typical risk factors for a reduced outcome in COVID-19 patients were more pronounced in patients requiring MV compared to non-MV patients. Surprisingly, the extension of these parameters could not distinguish between survivors and non-survivors in a mechanically ventilated subgroup.

These findings diverge remarkably from the findings, which were published before, analyzing generic COVID-19 cohorts. The differences in age-related mortality between different BMI-subgroups were a surprising observation. Many authors stress the negative effect of an increased BMI for the outcome of COVID-19 patients, partly even as a dose-response relationship (14-16). For our population, we cannot confirm this observation. Surprisingly, we found a strongly decreased risk of mortality for younger patients with an intermediate BMI. If we look at the relative mortality over all patients, this results in a U-shaped mortality risk. Interestingly, the nadir of the mortality curve seems not to lay in the “healthy” range of the BMI but in a slightly elevated range between 25 and 32, i.e. covering the preadiposity range. Holman et al. reported a quite similar distribution COVID-19 related mortality in a cohort of English diabetes patients (17). The causality of this finding remains unclear. Nevertheless, there are some hints that could support our results. In a large-scale study of the Global BMI Mortality Collaboration with more than 10 million analyzed participants, the risk of mortality caused by a respiratory disease, showed an U-shaped classifications with its nadir between 22.5 and 25 (18). So, also BMI values between 20 and 22.5 usually classified as normal weight (19), might have to be considered as underweight and thus disadvantageous. Underweight itself is known to be a risk factor in respiratory diseases and impairing pulmonary function (20-22). A very confusing result is the reduced mortality in the group of old patients with high BMI. According to other authors, these patients combine two highly endangering risk factors, but in our population, they show a clearly decreased mortality. Remarkably, in ARDS patients, outcome is better in patients with an increased BMI (23, 24). Therefore, the affection of the respiratory system, like negative effects of thoracic wall weight and abdominal fat mass on pulmonary compliance of obese patients, obviously does not lead to an increased mortality in COVID-19 patients. In the end, we have to acknowledge that the sample size was small, especially in this group. In general, mortality in the younger patient groups is increased indicating a tendency in contradiction to the epidemiological findings. In addition, an analysis of overall 89 covariates of comorbidities or clinical/diagnostic parameters did not explain the observed mortality profiles. These findings indicate that mortality within the ICU-patient cohort is mainly driven by covariates aside from BMI and age both playing a significant role in mortality factor analysis in generic COVID-19 cohorts. Further appropriate research considering the observed complexity regarding BMI and age needs to be carried out in bigger, but more selective populations.

Although mortality is influenced by BMI and age in a non-monotonic fashion, there are no significant differences in the length of MV among the different subgroups. The only remarkable finding is a trend for longer MV durations in survivors compared with non-survivors. This is not surprising, since patient data in part contain data from a specialized unit for weaning from MV. Some patients could not be weaned primarily after recovery from the acute COVID-19 disease and required treatment due to prolonged weaning, which results in long MV durations. Non-survivors, on the other hand, die 18 days after the onset of symptoms (25). Also, in another German COVID-19 cohort, patients had a particularly high probability of dying in the first 10 days of hospitalisation. In this cohort, length of stay of more than 18 days was associated with survival (12).

The present data possibly could give a hint that the disease evolution of severely ill COVID-19 patients includes a more complex interaction of risk factors and biometric variables. This indicates that the impact factors leading to either light or severe symptoms after COVID-19 infections may be different from those responsible for the evolution to death or recovery in severe cases. These hypotheses are supported by analyses of ICU patients without MV compared to MV patients. Between these two groups, we find the risk factors for a severe course of disease, which were described before, like increased laboratory parameters indicating an increased inflammatory level and coagulation status (26). Remarkably, these parameters were not able to discriminate between survivors and non-survivors as soon as MV had started. Additionally, the high correlation between all diagnostic baseline parameters and enrichment of comorbidities calculated between non-MV / MV patient cohorts as well as the lack of correlation between MV survivors / MV non-survivor cohorts indicate that the transition from non-MV to MV disease state is driven by mechanisms, which are less relevant for the following disease progression during MV. Also, with respect to duration of MV in survivors, there are signs indicating that the transition from non-MV to MV disease status is associated with mechanisms controlling severity of the disease, as assessed by the duration of MV. In contrast, the mechanisms controlling survival / non-survival within the MV subcohort show no correlation to duration of MV, supporting our hypothesis of the diversity of mechanisms controlling mortality and severity of disease.

Our study surely has limitations, which have to be considered. It has to be taken into account that the robustness of the analysis of the UKA cohort data is impaired by the small sample size, partially leading to small subgroups. To counteract this disadvantage, we used an approach for the analysis with variable group boundaries, which is based on the fuzzy logic concept. Yet, the small sample also prevented further examination on additional risk factors, like gender differences. Furthermore, the analyzed patients show an extraordinarily high severity of the disease and complex comorbidities, which required treatment in a university hospital making it difficult to transfer the results on other populations.

Nevertheless, we claim that the risk structures of the transitions from mild to severe disease states are structurally different to the risk structures within highly severe disease states. The analysis of pooled data from all disease states, which aims to investigate risk factors for mortality, reveals the convolution of risk profiles of both disease states, dominated by the critical step, namely the step from mild to severe stage. Hence, our findings from ICU patients may not be in contradiction to the results published from large, pooled studies.

From our retrospective study, we deduce the recommendation that statistical analysis of risk factors and epidemiological / therapeutic measures should be adapted to the apparently complex and diverse disease driving mechanisms also for bigger cohorts. We suggest that data analysis in COVID-19 patient cohorts should use methods that are able to find complex, non-monotonic multi-variate patterns, which are able to reflect mutual interference of parameters. This could infer the apparent complexity of the interference of disease evolution and recovery processes in critically ill COVID-19 patients.

## Conclusions

The aim of this study was to get a more detailed insight into the influence of biometric covariates on the outcome of COVID-19 patients with high degree of severity. We find that survival in mechanical ventilation is affected by complex interactions of covariates differing to the reported covariates associated with transition from mild to severe disease stages which are hidden in generic, non-stratified studies on risk factors. Hence, our study suggests that a detailed, multivariate pattern analysis on larger patient cohorts reflecting the specific disease stages might reveal more specific patterns of risk factors supporting individually adapted treatment strategies.

## Data Availability

The datasets of patients of University hospital RWTH Aachen, which were generated and analyzed during the current study, are not publicly available due to medical confidentiality but are available from the corresponding author on reasonable request.

## List of abbreviations

ARDS: Acute respiratory distress syndrome
BMI: Body mass index
COVID-19: Corona virus disease 2019
ECMO: Extracorporeal membrane oxygenation
ICU: Intensive care unit
MV: Mechanical ventilation
PCR: Polymerase chain reaction
PEEP: Positive end expiratory pressure
SARS-CoV2: Severe acute respiratory syndrome Coronavirus 2

## Declarations

## Ethics approval and consent to participate

This analysis was approved by the local ethical review board (EK 091/20; Ethics Committee, Faculty of Medicine, RWTH Aachen, Aachen, Germany).

## Consent for publication

Not applicable.

## Authors’ contributions

JB created the dataset of UKA patients and applied for consent of the local ethical review board. KS and AS analyzed the patient data and developed the prediction model. SF and JB interpreted the results from a medical perspective. SF, JB and AS wrote the manuscript. All authors read and approved the final manuscript.

## Acknowledgements

Not applicable.

## Authors’ information (optional)

Not applicable.

**Figure 1a:** Distribution of mortality across combined BMI – age data in COVID-19 ICU patient cohort **Figure 1b:** Distribution of length of MV across combined BMI – age data in COVID-19 ICU patient cohort **Figure 2:** Distribution of age for survivors/non-survivors stratified by BMI

**Figure 3a/b:** Mortality in BMI/age stratified subcohorts at 12 splitting levels reveals inhomogeneous, but non-monotonic distribution of mortality (3a). Mean mortality (weighted by subcohort size) indicate outliers of very low mortality in subcohort 3 and a slightly decreased mortality in subcohort 6. Labeling for BMI and age group: [L]: low, [M]: intermediate, [H]: high.

**Figure 4:** Length of mechanical ventilation is higher for survivors than for non-survivors in all subcohorts except cohort 3 (very low incidence of non-survival)

**Figure 5a/b:** There is a high correlation between significance levels of baseline diagnostic parameters between non-MV and MV patients (5a), but no correlation when comparing survivors and non-survivors in MV (5b).

## References

1. Xie Y, Wang Z, Liao H, Marley G, Wu D, Tang W. Epidemiologic, clinical, and laboratory findings of the COVID-19 in the current pandemic: systematic review and meta-analysis. BMC infectious diseases. 2020;20(1):640.

2. SARS-CoV-2 Steckbrief zur Coronavirus-Krankheit-2019 (COVID-19). Robert-Koch-Institut.. Accessed 30.10.2020.

3. Schilling J, Diercke M, Altmann D, Haas W, Buda S. Vorläufige Bewertung der Krankheitsschwere von COVID-19 in Deutschland basierend auf übermittelten Fällen gemäß Infektionsschutzgesetz. 2020.

4. Kluge S, Janssens U, Welte T, Weber-Carstens S, Marx G, Karagiannidis C. [Recommendations for critically ill patients with COVID-19]. Medizinische Klinik, Intensivmedizin und Notfallmedizin. 2020;115(3):175–7.

5. Palmieri L, Vanacore N, Donfrancesco C, Lo Noce C, Canevelli M, Punzo O, et al. Clinical Characteristics of Hospitalized Individuals Dying with COVID-19 by Age Group in Italy. The journals of gerontology Series A, Biological sciences and medical sciences. 2020.

6. Pastor-Barriuso R, Perez-Gomez B, Hernan MA, Perez-Olmeda M, Yotti R, Oteo J, et al. SARS-CoV-2 infection fatality risk in a nationwide seroepidemiological study. medRxiv. 2020:2020.08.06.20169722.

7. Salje H, Tran Kiem C, Lefrancq N, Courtejoie N, Bosetti P, Paireau J, et al. Estimating the burden of SARS-CoV-2 in France. Science. 2020;369(6500):208–11.

8. Wu Z, McGoogan JM. Characteristics of and Important Lessons From the Coronavirus Disease 2019 (COVID-19) Outbreak in China: Summary of a Report of 72?314 Cases From the Chinese Center for Disease Control and Prevention. JAMA. 2020;323(13):1239–42.

9. Patel U, Malik P, Usman MS, Mehta D, Sharma A, Malik FA, et al. Age-Adjusted Risk Factors Associated with Mortality and Mechanical Ventilation Utilization Amongst COVID-19 Hospitalizations-a Systematic Review and Meta-Analysis. SN comprehensive clinical medicine. 2020:1–10.

10. Zheng Z, Peng F, Xu B, Zhao J, Liu H, Peng J, et al. Risk factors of critical & mortal COVID-19 cases: A systematic literature review and meta-analysis. The Journal of infection. 2020;81(2):e16–e25.

11. Dreher M, Kersten A, Bickenbach J, Balfanz P, Hartmann B, Cornelissen C, et al. The Characteristics of 50 Hospitalized COVID-19 Patients With and Without ARDS. Deutsches Arzteblatt international. 2020;117(16):271–8.

12. Karagiannidis C, Mostert C, Hentschker C, Voshaar T, Malzahn J, Schillinger G, et al. Case characteristics, resource use, and outcomes of 10?021 patients with COVID-19 admitted to 920 German hospitals: an observational study. The Lancet Respiratory medicine. 2020.

13. Tang JW. COVID-19: interpreting scientific evidence – uncertainty, confusion and delays. BMC infectious diseases. 2020;20(1):653.

14. Cornejo-Pareja IM, Gómez-Pérez AM, Fernández-García JC, Barahona San Millan R, Aguilera Luque A, de Hollanda A, et al. Coronavirus disease 2019 (COVID-19) and obesity. Impact of obesity and its main comorbidities in the evolution of the disease. European eating disorders review: the journal of the Eating Disorders Association. 2020.

15. Du Y, Lv Y, Zha W, Zhou N, Hong X. Association of Body mass index (BMI) with Critical COVID-19 and in-hospital Mortality: a dose-response meta-analysis. Metabolism: clinical and experimental. 2020:154373.

16. Malik VS, Ravindra K, Attri SV, Bhadada SK, Singh M. Higher body mass index is an important risk factor in COVID-19 patients: a systematic review and meta-analysis. Environ Sci Pollut Res Int. 2020.

17. Holman N, Knighton P, Kar P, O’Keefe J, Curley M, Weaver A, et al. Risk factors for COVID-19-related mortality in people with type 1 and type 2 diabetes in England: a population-based cohort study. The lancet Diabetes & endocrinology. 2020;8(10):823–33.

18. Di Angelantonio E, Bhupathiraju SN, Wormser D, Gao P, Kaptoge S, de Gonzalez AB, et al. Body-mass index and all-cause mortality: individual-participant-data meta-analysis of 239 prospective studies in four continents. The Lancet. 2016;388(10046):776–86.

19. Obesity: preventing and managing the global epidemic. Report of a WHO consultation. World Health Organization technical report series. 2000;894:i-xii, 1-253.

20. Azad A, Zamani A. Lean body mass can predict lung function in underweight and normal weight sedentary female young adults. Tanaffos. 2014;13(2):20–6.

21. Do JG, Park C-H, Lee Y-T, Yoon KJ. Association between underweight and pulmonary function in 282,135 healthy adults: A cross-sectional study in Korean population. Scientific Reports. 2019;9(1):14308.

22. Moser J-AS, Galindo-Fraga A, Ortiz-Hernández AA, Gu W, Hunsberger S, Galán-Herrera J-F, et al. Underweight, overweight, and obesity as independent risk factors for hospitalization in adults and children from influenza and other respiratory viruses. Influenza and Other Respiratory Viruses. 2019;13(1):3–9.

23. O’Brien JM, Jr., Phillips GS, Ali NA, Lucarelli M, Marsh CB, Lemeshow S. Body mass index is independently associated with hospital mortality in mechanically ventilated adults with acute lung injury. Critical care medicine. 2006;34(3):738–44.

24. Zhi G, Xin W, Ying W, Guohong X, Shuying L. “Obesity Paradox” in Acute Respiratory Distress Syndrome: Asystematic Review and Meta-Analysis. PLoS One. 2016;11(9):e0163677.

25. Khalili M, Karamouzian M, Nasiri N, Javadi S, Mirzazadeh A, Sharifi H. Epidemiological characteristics of COVID-19: a systematic review and meta-analysis. Epidemiol Infect. 2020;148:e130–e.

26. Ponti G, Maccaferri M, Ruini C, Tomasi A, Ozben T. Biomarkers associated with COVID-19 disease progression. Critical reviews in clinical laboratory sciences. 2020;57(6):389–99.

