## Appendix for "Biometric covariates and outcome in COVID-19 patients: Are we looking close enough?"

**Influence of baseline values**

We checked differential baseline values across the survivors / non-survivors in each subcohort, compared to all survivors and non-survivors. Systematic differential expression in a subcohort compared to all MV patients are given in table 1.

Table 1: Baseline values, which are increased / decreased within the respective subcohort

| | | |
|---|---|---|
| Bilirubin | Decreased ($p < 0.05$) | Survivors [Low BMI/high age] |
| I:E | Increased ($p < 0.05$) | Survivors [Low BMI/high age] |
| Creatinine | Increased ($p < 0.05$) | Survivors [Low BMI/high age] |
| Tidal volume | Increased ($p < 0.05$) | Survivors [High BMI/high age] |
| $SpO_2$ | Increased ($p < 0.05$) | Non-Survivors [Intermediate BMI/low age] |

Surprisingly, the only significant difference in subcohort 3 is $SpO_2$ for Non-Survivors despite the highly significant deviation of mortality.

**Influence of comorbidities**

In analogy to differential levels of baseline diagnostic parameters, we assessed enrichment of comorbidities between survivors and non-survivors for each subcohort compared to survivors / non-survivors in all MV patients.

Comorbidities, which are enriched / suppressed within the survivor/ non-survivor populations of the 6 subcohorts are shown in table 2:

Table 2: Comorbidities, which are enriched / suppressed within the respective subcohort

| | | |
|---|---|---|
| Chronic Heart Failure | Enriched ($p < 0.05$) | Survivors [High BMI/low age] |
| Diabetes Mellitus | Enriched ($p < 0.01$) | Survivors [High BMI/low age] |
| Diseases of blood and blood forming organs | Suppressed ($p < 0.05$) | Non-survivors [Low BMI/high age] |
| Diseases of the digestive system | Suppressed ($p < 0.05$) | Non-survivors [Intermediate BMI/high age] |
| Diseases of the digestive system | Enriched ($p < 0.05$) | Non-survivors [High BMI/low age] |
| Sepsis | Suppressed ($p < 0.05$) | Non-survivors [Low BMI/high age] |

As the enrichment analysis didn’t result in neither highly significant enrichments of comorbidities nor difference in diagnostic baseline levels, corrections with respect to multiple testing show no robust significance. Hence, we cannot claim univariate explanations of the observed patterns in morbidity.

**Enrichment analysis of comorbidities in non-MV patients compared to MV patients**

Assessing the enrichment (Bonferroni-corrected $p < 0.01$) of comorbidities in MV-survivor / non-survivor population compared to non-MV population, we found comorbidities, which are strongly enriched in MV cohort (independent from survival) compared to non-MV patient cohort:

Table 3: Enrichment of comorbidities in MV population compared to non-MV population

| | **Non-MV** | **MV** |
|---|---|---|
| Diseases of the blood and blood-forming organs | log10(p) = -3.48 | log10(p) = -3.73 |
| Diseases of the genitourinary system | log10(p) = -2.62 | log10(p) = -3.98 |
| Diseases of the skin and subcutaneous tissue | log10(p) = -3.57 | log10(p) = -2.22 |
| Endocrine, nutritional and metabolic diseases | log10(p) = -3.83 | log10(p) = -3.3 |
| Symptoms, signs and abnormal clinical and laboratory finding | log10(p) = -3.92 | log10(p) = -3.98 |
| Sepsis | log10(p) = -4.8 | log10(p) = -6.3 |

Apparently, Sepsis is a syndrome, which is highly significantly enriched in MV-patient cohort compared to non-MV cohorts, both for survivors and non-survivors.
